# On the parameterization of mathematical models of infectious disease transmission structured by age at the start of the epidemic spread

**DOI:** 10.1101/2024.04.11.24305604

**Authors:** Santiago Sarratea, Gabriel Fabricius

**Affiliations:** Instituto de Investigaciones Fisicoquímicas Teóricas y Aplicadas (INIFTA), CONICET and Facultad de Ciencias Exactas, Universidad Nacional de La Plata, CC 16, Suc. 4, 1900 La Plata, Argentina; CCT CONICET La Plata, Consejo Nacional de Investigaciones Científicas y Técnicas, Argentina

## Abstract

Estimation of transmission and contact rate parameters among individuals in different age groups is a key point in the mathematical modeling of infectious disease transmission. Several approaches exist for this task but, given the complexity of the problem, the obtained values are always approximate estimations that hold in particular conditions. Our goal is to contribute to this task in the event of an emerging disease. We propose a methodology to estimate the contact rate parameters from the fraction of the incidence reported in each age group at the beginning of the epidemic spread. Working with an age-structured SIR model, we obtain an equation that relates the contact parameters to various epidemiological quantities that could be accessible through different sources. We apply the method to obtain information about the contact structure by age during the COVID-19 epidemic spread in Greater Buenos Aires (Argentina) in 2020. As we have the fractions of reported incidence by age but only rough estimations of other quantities involved in the method, we define several epidemiological scenarios based on various hypotheses. Using the different sets of contact parameters obtained, we evaluate control strategies and analyze the dependence of the results on our assumptions. The proposed method could be useful to obtain a fast first insight of a new emergent disease at the beginning of epidemic spread using the accessible information.

## 1 Introduction

Mathematical modeling has become a fundamental tool to study the transmission process of infectious diseases and to help in the design of control strategies for endemic and emergent diseases [1, 2]. In the design of mathematical models, assumptions about the network of social contacts through which the disease spreads play a central role. The characterization of this network is an extremely complex problem and highly dependent on the social conditions that exist at a given place and time. The advances that can be made in this characterization, based on the available epidemiological information, are one of the key points in the study of this problem. Depending on the available information, there is a variety of approaches to address this issue, from the simplest homogeneous mixing approximation, through approaches that involve different kinds of heterogeneities, to complex network or agent based models [3–5].

One of the first aspects usually considered to account for heterogeneity in the transmission process is the age of the individuals, since individuals of similar age usually have common social behaviors. For example, children go to school and often live with their parents, adults contact other adults at work, etc. Moreover, some diseases affect individuals of different ages in a different way, which justifies and enhances the relevance of this approach. Abundant epidemiological evidence of specific features observed in the transmission process of several diseases that depend on age has been reported [3]. In order to include the age dependence of the transmission process in compartmental mathematical models, *N*_*a*_ *× N*_*a*_ transmission matrices are usually defined, which account for the probability of transmission of infection among individuals in the different *N*_*a*_ age groups considered. Several methods, which involve different approximations, have been proposed to determine these matrices from epidemiological information [3, 6, 7]. In particular, in one of these methods, the 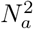 unknown matrix elements are reduced to *N*_*a*_, which are determined requiring that the model reproduces the forces of infections in the corresponding age groups obtained from epidemiological information at the endemic state [3, 8]. This reduction in the number of unknowns is performed forcing the transmission matrix to be symmetric and assuming a structure for it with N_a_ independent elements. This *structure* is contained in the so-called WAIFW (Who Acquires Infection from Whom) matrix [3]. Arbitrariness in the choice of the WAIFW structure that has to be constrained to only *N*_*a*_ elements is the main drawback of this method. Other approaches to the problem, specifically aimed at airborne infections, involve collecting information about individual mobility to generate synthetic populations [9] or focusing on data obtained from surveys about an individual’s daily conversations with other people [10]. In ref.[11], Wallinga et al. propose the “social contact hypothesis” that states that the number of potentially infectious contacts between individuals in two given age groups is proportional to the self-reported number of social contacts between them. This implies that all the properties of infectiousness and susceptibility of the pathogen transmission between individuals are assumed to be independent of age and accounted by a constant factor. Using information from a cross-sectional survey conducted in Utrecht (Netherlands) they conclude that their hypothesis gives reasonable results when checked against epidemiological data for mumps and influenza. Since then, many survey studies asking specific questions about daily contacts with other individuals were conducted throughout the world and this approach of estimating contact matrices has been widely used by mathematical modelers in the study of several infectious diseases [12, 13]. Although the social contact hypothesis does not consider the age dependence of the susceptibility to become infected with a given pathogen or the ability to transmit it, these elements could be explicitly incorporated if the information were available. For example, during the COVID-19 pandemic, Zhang et al. determined transmission matrices based on social contact ones but introducing age-dependent susceptibility [14]. Conversely, Franco et al. derived estimations of age-dependent susceptibilities and infectivities from reported incidences and contact matrices, which were estimated using survey data [15]. The main problem with the information extracted from conversational surveys lies in the definition of contact, which can have a different impact for different diseases, and in the need to do them at a certain time and place, given that their extrapolation to other social conditions is not obvious. In any case, given that none approach is entirely conclusive because of the complexity of the problem, a sensible practice (when possible) is to use several complementary approaches to confirm the predictions of the models [16–19].

In this work we propose a methodology with the purpose of connecting different model parameters with epidemiological quantities at the beginning of the epidemic spread. Working with an SIR model structured by age, an equation is obtained where the contact parameter matrix, the susceptibilities, infectivities, fractions of reported incidences and relative underreporting factors by age, as well as the reproductive ratio, *R*_*0*_, at the beginning of the epidemic spread, are connected. The method is applicable when the conditions are met for the total incidence to grow exponentially and the incidence fractions by age to remain constant within a certain time window. If the contact rate matrix is unknown, it is necessary to assume a structure for it with as many free elements as age groups are being considered, as in the case of the WAIFW structure.

As an example in which the methodology can be useful, we study the social contact structure during the onset of the COVID-19 epidemic in Greater Buenos Aires (GBA), Argentina, where there are no published survey-based estimates of contact matrices available. Between May and June 2020, there were specific conditions of restrictions, for example, there were no classes, social gatherings were suspended, and only certain essential worker areas (or others difficult to regulate) were operational. Under these conditions the epidemic spread with a total reported incidence growing at a sustained rate, while the fractions of the incidence in each age group stabilized for some time around somehow constant values. Using the available epidemiological information, we obtain plausible contact matrices for these specific conditions. We discuss various hypotheses about how the social contact structure was during that pandemic period and the effects of these hypotheses on the impact of vaccination control strategies, should a sterilizing vaccine had been available at that time. The analysis performed could be useful in the event of an emerging disease where mobility restrictions similar to those in place during the first wave of COVID-19 in Argentina were in effect.

## 2 Materials and Methods

Our method employs the conceptual framework of the Next Generation Matrix (NGM) approach, where equations of compartmental deterministic models are linearized [20, 21]. However, while most applications have been aimed at obtaining the basic reproductive number, *R*_*0*_ (which is the leading eigenvalue of the NGM), our development aligns more with applications seeking to relate the leading eigenvector of the NGM to incidences [15].

### 2.1 SIR model for an age-structured population

We consider an SIR model structured by age with *N*_*a*_ age groups defined by a given partition of ages: 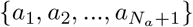. We are interested in considering the beginning of the epidemic development. So, given the narrow time window to be analyzed, we do not consider births or deaths, nor the transfers of individuals among age groups due to aging. Under these conditions, the dynamic evolution of the system is given by equations:

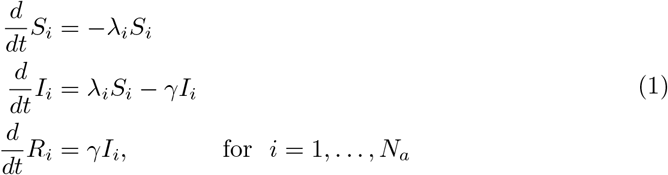

where *S*_*i*_, *I*_*i*_ and *R*_*i*_ are numbers of individuals in age group *i* that are susceptible, infected and recovered respectively. These are the dynamical variables of the model, where the sums *S*_*i*_ + *I*_*i*_ + *R*_*i*_ = *N*_*i*_ are constant in time from eq.(1). The force of infection *λ*_*i*_ is the rate at which susceptible individuals in age group *i* acquire infection and is given by:

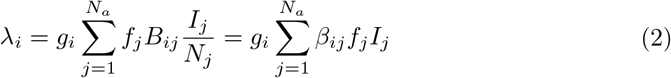

Here, *B*_*ij*_ is the rate of contacts that an individual of age group *i* has with individuals of age group *j*. The factor *I*_*j*_*/N*_*j*_ gives the probability that an individual in age group *j* is infected assuming that individuals within a given age group are uniformly mixed. The factors *g*_*i*_ and *f*_*j*_ are the susceptibility of an individual of age group *i* and the infectiousness of an individual in age group *j* respectively. They account for the ability of susceptible/infected individuals to become infectious/transmit infection, given a contact. Of course, *g*_*i*_ and *f*_*j*_ depend on the definition of contact. These age-dependent factors are introduced because for some diseases an age dependence of susceptibility and infectiousness has been reported (for example, for COVID-19, see refs.[22, 23]). We assume reciprocity of contacts, that is, that the total rate of contacts among individuals of age groups *i* and *j* is *B*_*ij*_*N*_*i*_ = *B*_*ji*_*N*_*j*_ [11]. Under this assumption, the contact matrix *β*_*ij*_ = *B*_*ij*_*/N*_*j*_ is symmetric. To avoid confusion, note that in ref.[3] (page 175) the notation *β*_*ij*_ is used to refer to the transmission matrix, which in our notation is *β*_*ij*_*g*_*i*_*f*_*j*_.

### 2.2 Start of the epidemic: linearization of the problem

We assume that at the beginning of the epidemic spread the number of infected individuals is very low in comparison with the total population, and that the amount of susceptible ones is large enough to be considered almost constant in time in spite of the infections that are being produced. That is,

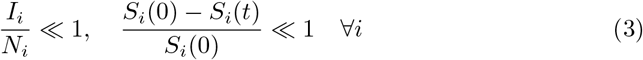

Then, in a time window (0, *t*_*s*_) such that reduction of the susceptible pool is not yet appreciable, Eq. (1) may be linearized, and the dynamic evolution of infected individuals approximated by:

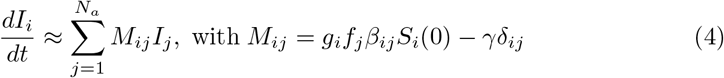

or in a more compact matrix form:

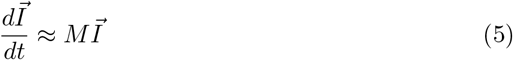

Under fairly general conditions[24], the solution to the above equation may be written as:

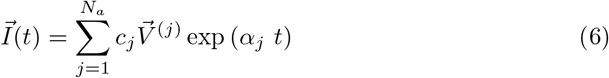

where *α*_*j*_ and 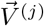 are the eigenvalues and eigenvectors of *M*, and *c*_*j*_ are constants determined from the initial condition:

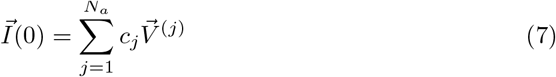

If there is a dominant eigenvalue *α*_*D*_, that is, an eigenvalue that is clearly higher than all the others, we may approximate 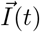 after a transient time, *t*_*trans*_, by

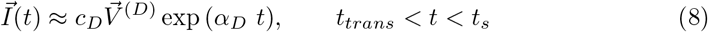

It can be proved that 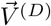 has all nonnegative components ^1^, as is necessary, since *I*_*i*_are positive numbers.

Let us denote *Y*_*i*_ as the incidence in age group *i*, that is, the rate at which new infections are produced at age group *i*. In the SIR model defined by Eq. (1):

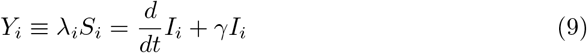

and when approximation (8) holds:

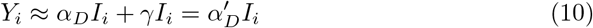

where we call 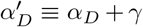 that is an eigenvalue of *M* + *γ𝕀* with the same eigenvector 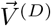, that is:

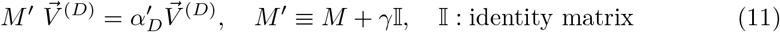

Then, the fraction of the incidence corresponding to age group *i, y*_*i*_, results:

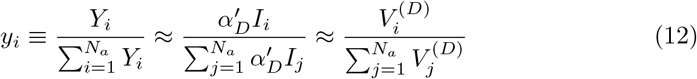

that is independent of time at this level of approximation, because all the components *I*_*i*_(*t*) depend on time through the factor exp (*α*_*D*_*t*) that is cancelled when computing the incidence fractions *y*_*i*_.

Note that the reproductive ratio, *R*_*0*_, defined as the number of new infections per infected individual produced in the system during an infectious period at the beginning of the epidemic is

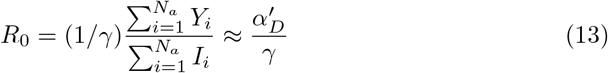

which is coherent with the definition of *R*_*0*_ as the largest eigenvalue of the Next Generation Matrix (NGM) that is *M* ^*′*^*/γ* for our system [20, 21].

### 2.3 The fraction of reported incidence by age

Our purpose is to use information about the relative fractions of reported incidences to parametrize the model. As is well known, there is underreporting of cases for all infectious diseases. And usually, this underreporting is dependent on age, according to the disease considered. For example, in the case of COVID-19 underreporting is expected to be lower for the elderly where the disease is more serious, and the opposite is expected in the case of pertussis, which is very serious for infants. Then, we assume that reported incidences are an age-dependent fraction of real incidences:

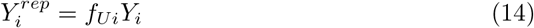

where *f*_*Ui*_ is the underreporting factor for the cases in agegroup *i*, which is expected to depend on *i*, but we assume it is independent of time in the narrow time window where Eq. (8) holds. Then, the fraction of reported incidences is

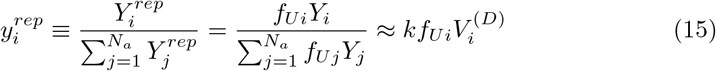

where *k* is a normalization constant independent of *i* and time. If we use Eq. (15) to write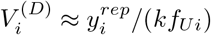, and Eq. (13) for 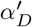 we may rewrite Eq. (11) as

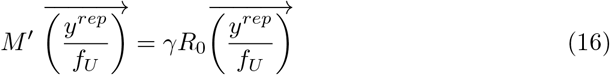

where the vector 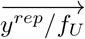 has components 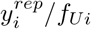, and matrix 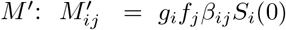. For simplicity we have written Eq. (16) as an equality and will consider it as such in what follows, keeping in mind that it is valid approximately and in the time window defined by Eq.(8). Under the assumptions we are working with, we do not expect the age-dependent underreporting to affect the calculation of *R*_*0*_ based on the reported data; because if the fraction of infected individuals by age group, *I*_*i*_, is also estimated from the reported data, it will be affected by the same underreporting factor *f*_*Ui*_. Then, using reported data to estimate *R*_*0*_:

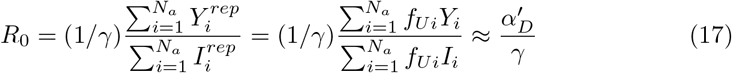

since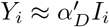. Finally note that Eq. (16) is not changed if the *f*_*Ui*_ are multiplied by a constant, which means that it is not necessary to know the absolute values of the underreporting factors but their relative values *f*_*Ui*_*/f*_*U1*_, for example.

### 2.4 Estimation of contact rate parameters *β*_*ij*_

Assuming *f*_*j*_, *g*_*i*_, 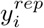, *f*_*Ui*_/(∑ *f*_*Uj*_), *γ*, and *R*_*0*_ are known, the *β*_*ij*_ could be obtained from (16) by reducing the number of unknowns from 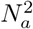 to *N*_*a*_. This can be done by defining a structure for matrix *β*_*ij*_ where there are only *N*_*a*_ unknown elements. An example of this type of structure is the WAIFW matrix that has been extensively used to obtain transmission matrix parameters from the knowledge of the forces of infection at the endemic state [3]. An example of WAIFW matrix for a case with 5 age groups that has been widely used in the literature [26] is given below:

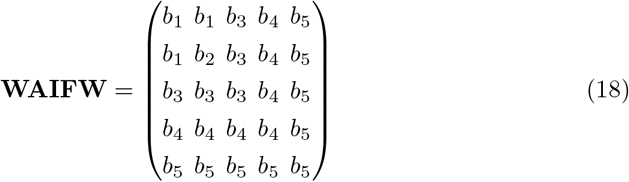

We will take for *β*_*ij*_ a more general form, that still depends linearly on *N*_*a*_ unknown elements:

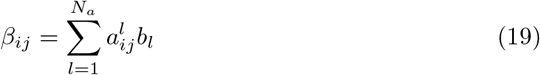

When this is introduced in Eq. (16), the unknown elements *b*_*l*_ can be obtained from the linear system:

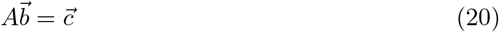

where:

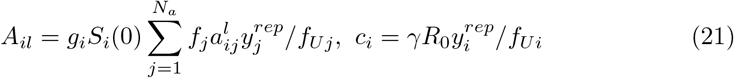

In the next section we give an example of this procedure for obtaining *β*_*ij*_ for the case of the COVID-19 epidemic spread in a region of Argentina.

But in fact, the proposed methodology has a broader scope. For example, it could happen that some (or all the) elements of *β*_*ij*_ were known from measurements of social contacts or from other sources, and that there were other unknown elements, for example, the *f*_*Ui*_. If this was the case, Eq. (16) would provide a useful tool to work with to estimate the missing parameters.

## 3 Application: the initial growth of COVID-19 in Argentina

In this section we apply the method developed in Section 2 to the initial spread of COVID-19 in 2020 in the Greater Buenos Aires (Argentina) with the purpose of obtaining information about social contacts at that time and place. In Argentina (and particularly in the GBA) there are no published measurements or estimates of social contacts between people, neither before, during nor after the pandemic.

We assume that between May 20 and June 10 (approximately), there are appropriate conditions to apply the method discussed in Section 2. Figure 1 shows the total incidence at the beginning of the epidemic and the fractions of reported incidences in different age groups, 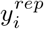. The 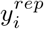 are seen to oscillate at first but eventually level off and remain approximately constant in the chosen period of time, which is consistent with the hypothesis that there is a dominant eigenvalue of the matrix *M*. The constancy of the 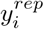 is also consistent with the relative underreporting factors remaining more or less constant during that period. It is worth noting that there were no changes in testing policies during that period either[27]. Now we give arguments to support the hypothesis that the social conditions remained more or less constant in that period, which justifies assuming that social contacts between groups were more or less well defined. The first confirmed case of COVID-19 in Argentina was on March 3, 2020, and on March 20 a very restrictive lockdown was decreed. Although most of the measures (including the suspension of school activities) were officially maintained in the GBA until the beginning of November 2020, in practice they were gradually relaxed, especially those related with to work and commercial activities. This relaxation, which could be observed in the mobility data from Google [28] (see Fig.S1a from ref.[29]), was sharp in the first two months and much softer in the following months. This justifies assuming that the β_ij_ remained more or less constant between May 20 and June 10. Finally, the total cases reported from the beginning of the epidemic until May 20 represent a fraction of 0.00045 of the GBA population, so, even when underreporting was high, it is a good approximation considering the whole population as susceptible when the methodology is applied.

**Fig. 1:**
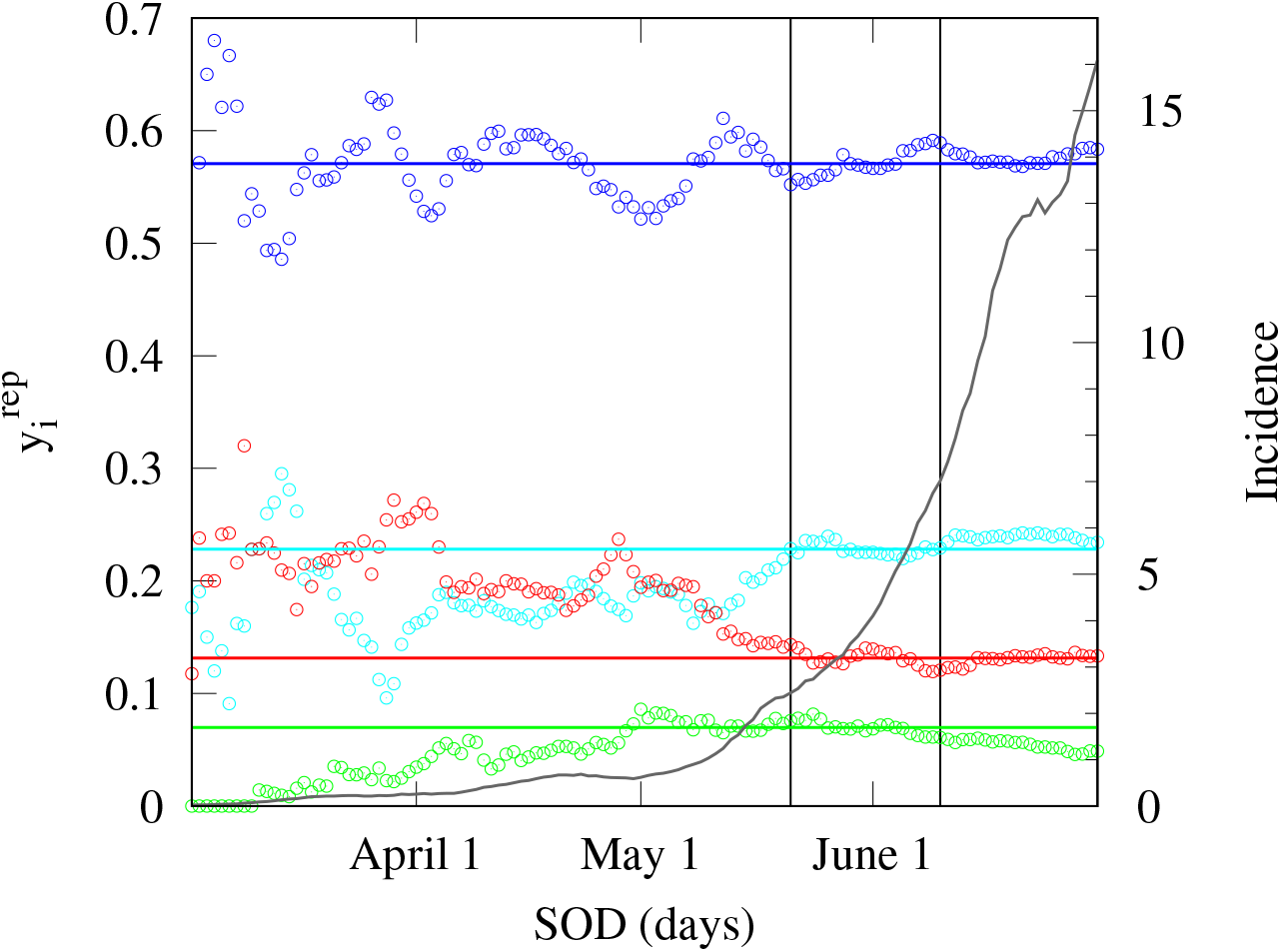
COVID-19 reported incidence in Greater Buenos Aires between March and July 2020. Total reported incidence in daily cases per 100,000 inhabitants (grey line, right y-axis) and fraction of reported incidence by age (colored circles, left y-axis), 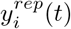, as a function of the symptom onset date (SOD). The four age groups, *i*, considered are: 0-15y (green), 15-30y (cyan), 30-60y (blue), and greater than 60y (red). The horizontal colored lines are the mean values of 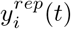 in the time window defined by the dates May 20 and June 10 indicated in the figure with vertical black lines. The curves are constructed with data taken from ref.[30] and smeared with a square window of a week. Details about data processing are given in ref. [29].

We have chosen four age groups in our study: 0-15y, 15-30y, 30-60y, and greater than 60y considering that each one of those groups had some common behavior features. The 0-15y group had low activity and basically at home, the 15-30y group had some social activity (informal sports, since formal activities were prohibited, and social gatherings) and some individuals in this group already worked. The 30-60y group is where the labor force is most concentrated and has probably been the most active. The group over 60 years was the riskiest and there was persistent insistence that they respected the restrictions by staying at home.

It should be mentioned that we have also analyzed the fractions of reported incidences taking smaller groups: 0-10y, 10-20y, 20-30y, 30-40y …, and the same constancy of the 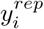 is observed, which would allow applying the method, but the number of unknowns to be determined would be much higher and we prefer to work with a reduced number of groups.

### 3.1 Model parameters and epidemiological scenarios

We take *N*_1_ + *N*_2_ + *N*_3_ + *N*_4_ = 1, so, *N*_*i*_ represents the fraction of the population in age group *i*, and *S*_*i*_ the fraction of the population in this group that is susceptible. We assume *S*_*i*_(0) = *N*_*i*_, and take for *N*_*i*_ the corresponding fractions of the population in each age group from the census carried out in Argentina in 2010 [31]: 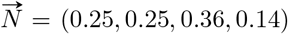. For the 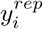 we take the mean values of the fractions of reported incidence by age between May 20 and June 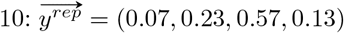. For *R*_*0*_ in this period we take the value 1.5, estimated in ref.[29] using an SEEIIR stochastic model to analyze reported data in GBA. The same estimation is obtained using an SIR deterministic model with *γ* = 1/(10 days) to fit the total incidence of Fig.1 between May 20 and June 10. Beyond having to assume a *structure* for *β*_*ij*_ matrix, we also have no accurate estimates of the susceptibilities *g*_*i*_, infectivities *f*_*i*_ and underreporting factors *f*_*Ui*_. Then, in the following sections, we perform some assumptions to estimate these magnitudes and define different scenarios compatible with the available epidemiological information. For *g*_*i*_ and *f*_*i*_, as we will see in Subsection 3.1.2, at most there are estimations of the relative values of these parameters between children and adults. So, to simplify the analysis, in the following we call *g*_*i*_ and *f*_*i*_ the relative values of the susceptibilities and infectivities taking max(*g*_*i*_)=max(*f*_*i*_)=1 and we modify the meaning of *β*_*ij*_ that will absorb these constants. That is,

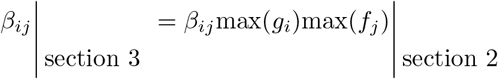

So, in the present section, the *β*_*ij*_ do not represent only contacts but include a constant factor (independent of *i* and *j*) related to the transmissibility of the infectious process that depends on each disease. However, the relative values between the different elements of *β*_*ij*_ are the same as those between the contact matrix of Section 2.

#### 3.1.1 Structure of contact matrices

We consider two cases: CM1 and CM2. The CM1 proposal explicitly assumes that the 30-60y age group is the one that drives the contagion process (see Table 1). Based on previous considerations, we assume that individuals in 0-15y age group remain mostly at home and the contagion occurred basically through contact with parents, so we take *β*_1*j*_ = 0, except for *j* = 3. Of course, there can be contact between siblings, but we assume that it can be taken into account effectively through *β*_13_, since the source of infection is the parents. We consider that an individual in age group 15-30y has the same probability of contacting one in 30-60y as one in 0-15y (*β*_23_=*β*_13_), but contacts with other individuals in the same age group are allowed in this case. We consider that the elderly are confined and only have contact with their children who provide them with food and care. Table 1 also shows the matrix structure proposed for the CM2 case, which is based on a somehow different hypothesis, trying to force some assortative character. Here we consider that children have greater contact with each other (assuming they contact other children in the neighborhood) and, to a lesser extent, with groups 2 and 3. We assume that individuals in age groups 15-30y and 30-60y may contact in outdoor activities and propose *β*_23_ is proportional to the average of the level of activity that individuals have in their own age groups. In fact, it is half of the average to induce assortativeness. Older people contact people between 30 and 60 years old but also, to a lesser extent, younger people. We continue (as in CM1 structure) assuming that the interaction between the elderly can be neglected and we take *β*_44_ = 0, but we have done tests modifying this matrix and taking *β*_44_ = *b*_4_, and the analysis is not qualitatively modified.

**Table 1:** Matrix structures used in Section 3. Each structure, CM1 and CM2, defines the 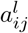 for the corresponfding contact matrix *β*_*ij*_ in Eq. (19).

| CM1 | CM2 |
| --- | --- |
| $\begin{pmatrix} 0 & 0 & b_1 & 0 \\ 0 & b_2 & b_1 & 0 \\ b_1 & b_1 & b_3 & b_4 \\ 0 & 0 & b_4 & 0 \end{pmatrix}$ | $\begin{pmatrix} b_1 & b_1/2 & b_1/2 & b_4/4 \\ b_1/2 & b_2 & (b_2 + b_3)/4 & b_4/2 \\ b_1/2 & (b_2 + b_3)/4 & b_3 & b_4 \\ b_4/4 & b_4/2 & b_4 & 0 \end{pmatrix}$ |

#### 3.1.2 Relative susceptibilities and infectivities

It is very complex to determine the dependence of susceptibility and infectivity on age in the case of COVID-19 because many biological, environmental, and epidemiological factors are involved in studies on the subject, in addition to the fact that since children often have milder symptoms, many cases go unnoticed [22]. However, there appears to be some evidence that leans in the direction of lower susceptibility and infectivity in children. Davies et al., using mathematical modelling to analyze epidemiological data, estimate that susceptibility in 0-20y age group is about half that of adults [32]. In a review that includes 32 studies in their analysis, Viner et al. obtained that, on average, susceptibility for 0-20y age group is about 0.56 that of adults, and for 0-15y age group, somehow less[23]. However, the analysis shows a great heterogeneity in the results obtained in the different studies. For example, in a study of 391 cases and their contacts in Shenzhen (China) the authors found that children were as likely to be infected as adults [33]. Concerning infectivity, information is even scarcer. In a review that includes several studies in schools it was found that lower infection attack rates were reported in students compared to school staff [34]. In another study performed in households where a stochastic dynamic model was used and the relative infectivity between children and adults was considered as an explicit variable of the model, an infectivity value of 0.63 was obtained [35]. So, in the present work we have considered two epidemiological scenarios for relative susceptibility and infectivity in the different age groups, the scenario gf0 where *g*_*i*_ and *f*_*i*_ are all equal: 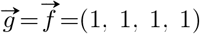, and the scenario gf1 where *g*_*i*_ and *f*_*i*_ have reduced values for children: 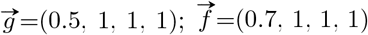).

#### 3.1.3 Relative underreporting

The relative underreporting among different age groups could be obtained, for example, by comparing the reported case numbers with age-discriminated serological studies. Since we do not have this information for the Greater Buenos Aires area in Argentina, we will define scenarios based on the qualitative information we have. It is reasonable to assume that the elderly report the most because, given that they have the most severe and symptomatic form of the disease, they are easier to detect, and also more attention is paid to them. Based on opposing arguments, it is sensible to assume that children report the least. Three scenarios have been considered: fU0: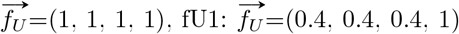, and 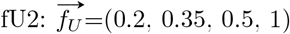. A priori, we consider a scenario when all the *f*_*Ui*_ are not equal to be more plausible for the reasons mentioned. The scenario fU0 could be feasible in a place where serology had been conducted along with intensive contact tracing, which was not the case in the Greater Buenos Aires area. However, we introduce it for the purpose of having a reference for comparative purposes. We have also considered scenarios with 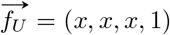, with 0.1 < *x* < 0.4, but this only introduces minor changes in the results obtained for fU1, which are discussed at the end of Subsection 3.2. Finally, the comparison with estimates of contact parameters based on surveys led us to consider two additional underreporting scenarios, fU3 and fU4, which are described in Section 3.3.

### 3.2 Results

#### 3.2.1 Determination of the contact parameters

Table 2 summarizes the age-dependent parameters discussed in the previous section that, together with the assumed value of *R*_*0*_ = 1.5, and the contact structures of Table 1, allow us to obtain the *β*_*ij*_. For simplicity, we will adopt the unit of time as 1/*γ*, consequently, in the subsequent expressions, the *β*_*ij*_ values will be in units of *γ*, and we will omit *γ* from the formulas.

**Table 2:** Parameters used to determine the contact rates, *β*_*ij*_, for the different epidemiological scenarios. Fractions of reported incidences, 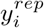, fractions of population, *N*_*i*_, suceptibilities, *g*_*i*_, infectivities, *f*_*i*_, and underreporting factors, *f*_*Ui*_, for each age group *i*. The last column lists the code to denote a possible set of parameter values or the reference of the data used in the case of fixed parameters. For *R*_*0*_ we have taken 1.5 (see Section 3.1).

| $i \equiv$ age group | 1 | 2 | 3 | 4 | |
| --- | --- | --- | --- | --- | --- |
| age interval | 0-15 | 15-30 | 30-60 | >60 |  |
| $y_i^{rep}$ | 0.07 | 0.23 | 0.57 | 0.13 | [30] |
| $N_i$ | 0.25 | 0.25 | 0.36 | 0.14 | [31] |
| $g_i$ | 1.0 | 1.0 | 1.0 | 1.0 | gf0 |
| $f_i$ | 1.0 | 1.0 | 1.0 | 1.0 | |
| $g_i$ | 0.5 | 1.0 | 1.0 | 1.0 | gf1 |
| $f_i$ | 0.7 | 1.0 | 1.0 | 1.0 | |
| $fU_i$ | 1.0 | 1.0 | 1.0 | 1.0 | fU0 |
| $fU_i$ | 0.4 | 0.4 | 0.4 | 1.0 | fU1 |
| $fU_i$ | 0.2 | 0.35 | 0.5 | 1.0 | fU2 |

Each epidemiological scenario is defined by the choice of a matrix structure (CM1 or CM2), a set of susceptibilities and infectivities (gf0 or gf1), and the underreporting factors (fU1, fU2, or fU3). The twelve scenarios obtained will be denoted by: CM1-gf0-fU1, CM1-gf0-fU2, …, etc. In Table 3 are shown the values of the *b*_*i*_ obtained for each scenario through Eqs. (20) and (21) and in Table 4 the resulting matrices. We begin by analyzing the values obtained for CM1-matrix structure. From inspection of the first and fourth rows of Table 3 it does not seem very plausible that *β*_33_ = b_3_ is of the same order as *β*_34_ = *b*_4_, since the elderly are assumed to be mostly confined, as opposed to the 30-60y group. In this sense, the fU1-scenarios (2nd and 5th rows) give more plausible values for *b*_3_ and *b*_4_, suggesting that the underreporting was indeed greater for the 30-60y group. Note that for these cases, the value 0.4 proposed for *f*_*Ui*_(*i ≠* 4) in fU1 only affects the relation *b*_4_/*b*_3_, so, other values such as 0.3 or 0.5 are also plausible ones. The negative value obtained for *b*_2_ in CM1-gf1-fU2 scenario indicates that there is something wrong. The problem is that assumption fU2 implies much greater actual incidence of children than what was reported and the reduced susceptibility and infectivity assumed for children in gf1 implies more activity (contacts) of children for a given transmission. This leads to a large value for *b*_1_ that, for case CM1, also implies a large contribution to 15-30y incidence that cannot be matched to reported data.

**Table 3:** Constants *b*_*i*_ that determine the matrix contact rates *β*_*ij*_ for the matrix structures of Table 1 in different epidemiological scenarios. The upper part of the table contains the values for the six CM1-scenarios and the lower part for the six CM2-scenarios. The *b*_*i*_ are in units of *γ* (1/*γ* is the mean duration of infection). The matrices for the twelve scenarios are shown in Table 4

| scenario | $b_1$ | $b_2$ | $b_3$ | $b_4$ |
| --- | --- | --- | --- | --- |
| CM1-gf0-fU0 | 0.73 | 4.21 | 3.22 | 2.45 |
| -fU1 | 0.73 | 4.21 | 3.69 | 0.98 |
| -fU2 | 1.83 | 2.83 | 2.41 | 1.22 |
| -gf1-fU0 | 1.47 | 2.37 | 2.89 | 2.45 |
| -fU1 | 1.47 | 2.37 | 3.36 | 0.98 |
| -fU2 | 3.67 | -0.38 | 1.14 | 1.22 |
| CM2-gf0-fU0 | 0.75 | 2.09 | 3.14 | 1.99 |
| -fU1 | 0.87 | 2.25 | 3.46 | 0.80 |
| -fU2 | 1.65 | 2.93 | 2.96 | 0.90 |
| -gf1-fU0 | 1.72 | 2.05 | 3.11 | 2.01 |
| -fU1 | 1.84 | 2.21 | 3.44 | 0.80 |
| -fU2 | 3.64 | 2.79 | 2.86 | 0.91 |

**Table 4:**
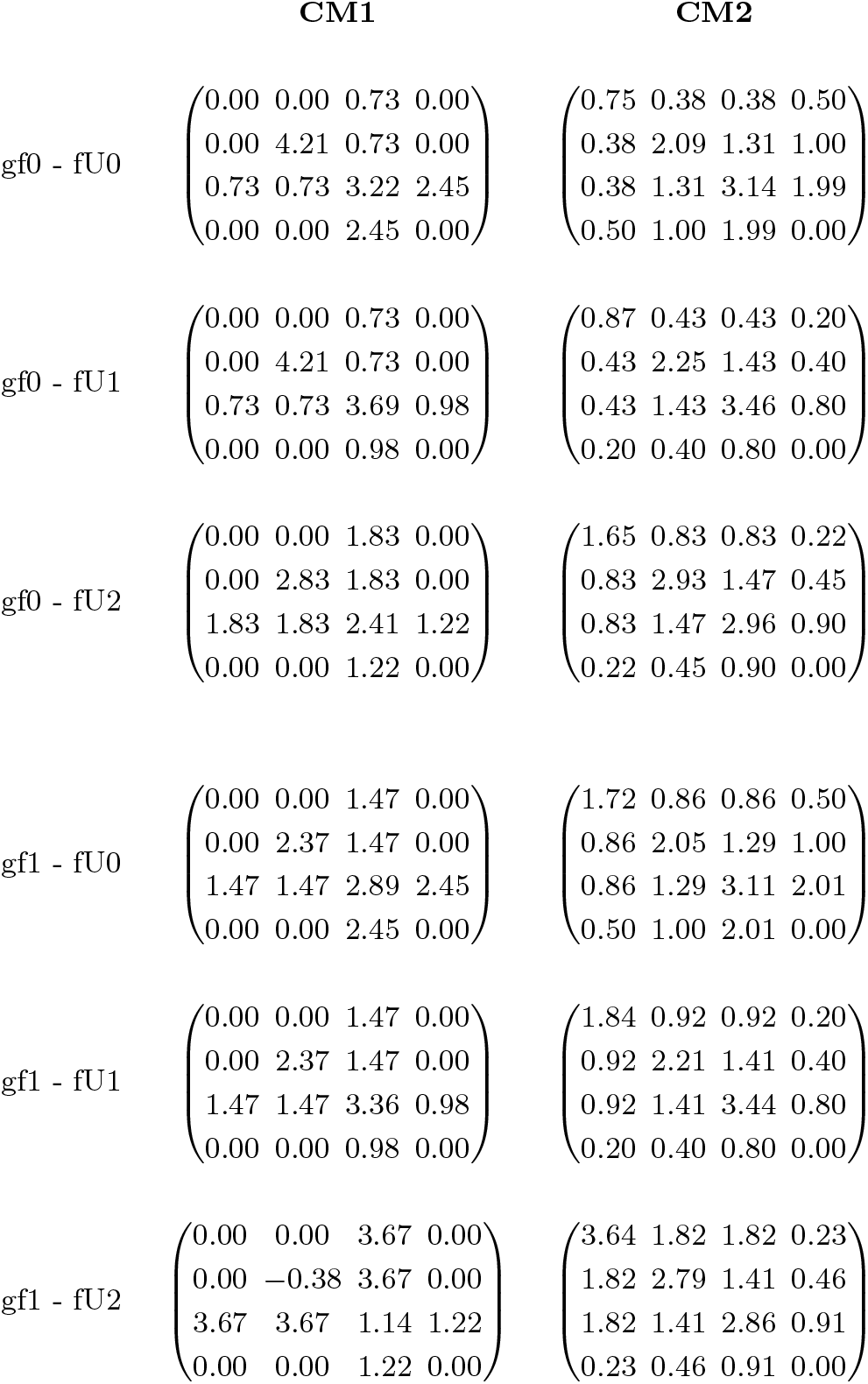
Contact matrices *β*_*ij*_ (in units of *γ*) for the twelve epidemiological scenarios considered in Section 3.

The scenarios corresponding to CM2-matrix structure are simpler to analyze looking at the matrices in Table 4. It can be seen that even when the proposed structure for CM2 is quite different than CM1-structure, the same qualitative considerations already made for CM1 are still valid in the case of CM2-structure. For example, in the case of CM2-gf1-fU2 (even when nonnegative values are obtained) the children have much more contacts than expected compared to the other age groups for a situation with no school and reduced related activities. However, CM2 and CM1 matrices are different, and this has some consequences on the control strategies analyzed in the next section.

From the results presented in the present section, scenarios gf1-fU2 do not seem plausible. The fU0 scenarios are not very plausible either, because the elderly end up with too many contacts given the restrictions that were imposed on them. Then, we will focus the following analysis on the fU1 and gf0-fU2 scenarios. However, it is important to consider that there are intermediate possibilities between fU0 and fU1 that could be feasible. Therefore, we will present results with fU0 with the aim of observing how the results of fU1 would be modified if 0.4 < *f*_*Ui*_ < 1 for *i* ∈ (0, 3).

#### 3.2.2 Vaccination strategies to stop transmission

In this section we analyze different control strategies assuming that a vaccine that protected against transmission existed when the pandemic started. Of course, this was not the case, and the pandemic is over, but the idea is to give an example of the usefulness of the matrices obtained in the previous section. So we discuss, with the restriction conditions that existed in the period studied (with R0 around 1.5), the relative advantages of different vaccination strategies with the purpose of halting transmission. Although campaigns may be centered on other objectives, such as reducing mortality or hospitalization. For more comprehensive studies on vaccine allocation based on specific objectives see, for example, the work by Miura et al. ref.[36].

If the vaccine has an effectiveness VE and a proportion *p* of the population is vaccinated, the immunized fraction is *p*VE. For simplicity, in what follows, we will assume VE=1 (100% effectiveness), which is not a realistic situation but simplifies the analysis. The largest eigenvalue of the NGM, usually denoted as *ρ*(NGM), has the meaning of a threshold parameter: if its value is less than 1, the disease is not able to produce an epidemic [20, 21]. In our problem,

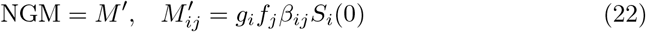

with *β*_*ij*_ in units of *γ*. When the whole population is susceptible *S*_*i*_(0) = *N*_*i*_ and *ρ*(NGM)=*R*_0_=1.5. When a fraction *p*_*i*_ of individuals in age group *i* are immunized, *S*_*i*_(0) = (1 − *p*_*i*_)*N*_*i*_, and the elements of the NGM are given by:

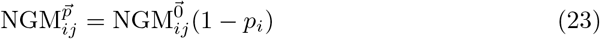

where 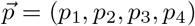 and 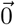 is the null vector. There are different possible choices of 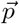 to achieve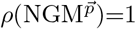. In particular, when the same fraction of individuals *φ* are immunized in each age group,

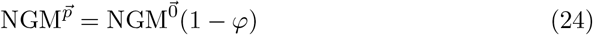

so, 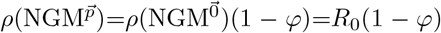, and the condition to stop transmission leads to the classical result *φ* = 1 *−* 1/*R*_0_ [3].

In Table 5 are shown the results obtained when immunizing with four vaccination strategies for different epidemiological scenarios. We first focus on the more plausible fU1 scenarios and consider strategy (i), where only group 3 is immunized. As can be seen in Table 5, except for scenario CM1-gf0, it is always possible to stop transmission immunizing a fraction of around 0.45 of group 3. It means that around 16% of the total population should be immunized. If vaccination were applied homogeneously, approximately twice as many vaccines would be needed. This result is quite independent of the relation *f*_*Ui*_/*f*_*U*4_=0.4 (for *i≠* 4) assumed for scenario fU1 because, as can be seen in the table, results for fU0 scenarios (where the underreporting for all age groups is the same) are very similar for this strategy. While many vaccines are saved with this strategy, the at-risk population would remain exposed, which would entail a risk in case the objective of halting transmission is not achieved. So, we have considered the strategy of immunizing 100% of the at-risk group and, under those conditions, determining the critical fraction of the 30-60y age group to be immunized. In this case, there is indeed a difference between the fU0 and fU1 scenarios because in the fU0 scenarios, the relative contribution of the elderly to transmission is higher. Therefore, with 100% of the elderly individuals immunized, it is necessary to immunize a smaller number of individuals in the 30-60y age group to halt transmission. This strategy, which is less risky, requires a greater quantity of vaccines, but always less than the 33% that would be necessary if immunization were done homogeneously. The strategy of immunizing groups 1, 2, and 3 (which includes all individuals < 60y) does require a significantly larger quantity of vaccines, but it still remains lower than if vaccination were done homogeneously. Only when this strategy is complemented as before by vaccinating 100% of the elderly, more vaccines are required than when vaccinating the entire population homogeneously.

**Table 5:** Vaccination strategies to stop transmission in different epidemiological scenarios. The vector 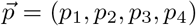 indicates the fraction *p*_*i*_ immunized in each age group *i* for each strategy. In each case the critical fraction *φ* of the corresponding agegroup(s) to be immunized is determined requiring *ρ*(NGM)=1. In cases where a dash is assigned instead of a number, it means that it is not possible to stop the transmission with that strategy. The percentage of the whole population immunized in each case is given by vacc=100 ∑_*i*_ *p*_*i*_*N*_*i*_.

| Scenario | Strategy |  |  |  |  |  |  |  |
| --- | --- | --- | --- | --- | --- | --- | --- | --- |
|  | (i) |  | (ii) |  | (iii) |  | (iv) |  |
| | $\vec{p}=(0, 0, \varphi, 0)$ | | $\vec{p}=(0, 0, \varphi, 1)$ | | $\vec{p}=(\varphi, \varphi, \varphi, 0)$ | | $\vec{p}=(\varphi, \varphi, \varphi, 1)$ | |
| | $\varphi$ | vacc | $\varphi$ | vacc | $\varphi$ | vacc | $\varphi$ | vacc |
| CM1-gf0-fU1 | — | — | — | — | 0.34 | 29.1% | 0.32 | 41.7% |
| CM2-gf0-fU1 | 0.45 | 16.1% | 0.42 | 29.3% | 0.34 | 29.1% | 0.32 | 41.7% |
| CM1-gf1-fU1 | 0.44 | 16.0% | 0.43 | 29.5% | 0.34 | 29.1% | 0.32 | 41.7% |
| CM2-gf1-fU1 | 0.45 | 16.2% | 0.43 | 29.4% | 0.34 | 29.1% | 0.32 | 41.7% |
| CM1-gf0-fU0 | — | — | — | — | 0.37 | 31.7% | 0.26 | 36.4% |
| CM2-gf0-fU0 | 0.48 | 17.4% | 0.33 | 25.9% | 0.37 | 31.6% | 0.25 | 35.8% |
| CM1-gf1-fU0 | 0.47 | 16.9% | 0.37 | 27.2% | 0.37 | 31.6% | 0.26 | 36.1% |
| CM2-gf1-fU0 | 0.48 | 17.4% | 0.33 | 26.1% | 0.37 | 31.6% | 0.25 | 35.8% |
| CM1-gf0-fU2 | 0.56 | 20.1% | 0.54 | 33.6% | 0.34 | 29.2% | 0.32 | 41.5% |
| CM2-gf0-fU2 | 0.64 | 23.2% | 0.61 | 36.0% | 0.34 | 29.2% | 0.32 | 41.5% |

Similar qualitative considerations arise when scenarios gf0-fU2 are considered, but in these scenarios the higher underreporting assumed for groups 1 and 2 with respect to group 3 leads to a higher weight of these groups in transmission compared with fU1 scenarios. So, concerning the strategy where only group 3 is vaccinated, a higher fraction of this group should be immunized to stop transmission.

Finally, it should be mentioned that when scenarios with underreporting factors given by 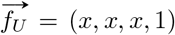, with 0.1 *< x <* 0.4, are considered, the results obtained are almost identical to those presented in Table 5 for the fU1 scenario. This happens because for *x* = 0.4, the contribution of the elderly to transmission is already sufficiently low, so the results no longer change if *x* decreases further.

### 3.3 Comparison with contact parameter estimates derived from survey data

As previously stated, there are no publicly available survey-based estimates of social contacts in Argentina. The extrapolation of contact measurements carried out at a specific time, place, and under certain mobility conditions to a situation with different characteristics is a very delicate matter. We conducted a comprehensive literature search for survey studies carried out during the COVID-19 pandemic but did not find any that specifically mirrored the conditions observed in Greater Buenos Aires (GBA) during the studied period. These conditions included the suspension of classes across all educational levels, while the epidemic spread with an *R* value clearly greater than 1.

During the COVID-19 pandemic, some authors used survey data collected before the pandemic to infer the impact of control measures on contact matrices during the pandemic [37, 38]. Since survey studies generally discriminate the location where the contact occurred, the matrix *B*_*ij*_ can be obtained as the sum of different contributions, for example,

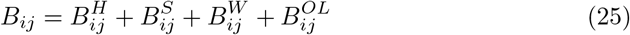

where 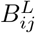 is the average number of contacts that an individual of age group *i* has with individuals of age group *j* at location *L* (*H* stands for home, *S* for school, *W* for work, and *OL* for “other locations” as transport, leisure, etc.). The discrimination of contacts into different contributions could be used to make a rough estimation of the resulting matrix, 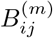, when restrictive measures associated with specific spaces have been implemented:

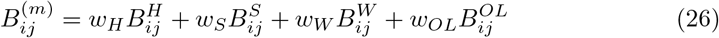

where restrictions are taken into account through factors *w*_*L*_ assigning weights to contacts in location *L* (for example, *w*_*S*_ = 0 would imply a total suspension of classes). We state that 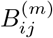 constitutes a rough estimate of the actual contacts that exist in a context of restrictions, recognizing that alterations in contacts within one setting are likely interconnected with changes in another setting. For example, the suspension of classes undoubtedly leads to adjustments in contacts among students in various locations. Much cruder and less reliable (according to the authors of the present work) are the extrapolations that attempt to derive the structure of social contacts in one country from contact surveys conducted in another, incorporating specific population, school-related, and other statistical indices [39]. Moreover, in cases like Argentina, where habits and social characteristics vary significantly across different regions, the meaning and applicability of the contact matrix obtained for the entire country are not clear.

Having outlined some of the troubles and considerations associated with utilizing indirect information from survey studies, we now present an estimation of this kind with the aim of analyzing the contact matrices yielded by this approach and comparing with the ones obtained through our method. We have taken contact measures conducted in conditions without restrictions from the widely used POLYMOD study [12] for three countries (Netherlands, United Kingdom and Italy) and also the synthetic matrices proposed by Prem et al. projected for Argentina [39]. These matrices are expressed in 5-year age groups, so we performed the following transformation to obtain matrices in each location expressed in the age groups we have considered in our study [40]:

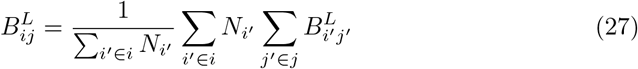

where for *i*=1 (0-15y age group), for example, the index *i*^*′*^ runs for age groups 0-5y, 5-10y and 10-15y, and *N*_*i*_^*′*^ is the population in age group *i*^*′*^. We used equation (26) taking *w*_*H*_=1, *w*_*S*_=0, *w*_*W*_ =0.5, and *w*_*OL*_=0.25. Neither the POLYMOD matrices [12] nor the synthetic matrices from ref.[39] satisfy the reciprocity condition that we have imposed in our study. As this can lead to spurious results [41], we transform the matrices 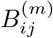 to satisfy reciprocity as follows [38]:

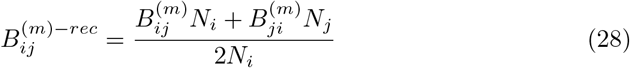

where *N*_*i*_ is the population of age group *i* in the corresponding country. Finally, we obtained the estimated symmetric contact matrices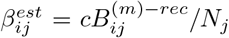, where the constant *c* is determined to ensure that the largest eigenvalue of the corresponding NGM: 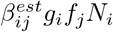 is 1.5 taking *g*_*i*_ and *f*_*j*_ according to gf1 scenario ^2^. In Table 6, the four obtained matrices are shown. A common feature in these matrices, not observed in those obtained with our method for gf1 (presented in Table 4), is that *β*_22_ > *β*_33_. In fact, *β*_22_ is the largest element in the entire matrix (except for the case of POL-NL). Another noteworthy characteristic is the similarity between the POL-IT and SYN-ARG matrices.

**Table 6:** Contact matrices 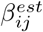 (in units of γ) estimated using survey data from the POLY-MOD study [12] for the Netherlands (POL-ND), Italy (POL-IT), and the United Kingdom (POL-UK), and using synthetic matrices projected for Argentina from Prem et al. [39] (SYN-ARG). The indices *i, j*=1,…4 correspond to age groups 0-15y, 15-30y, 30-60y, and greater than 60 years. The estimation procedure, described in Section 3.3, involves assigning weights *w*_*H*_=1, *w*_*S*_=0, *w*_*W*_ =0.5, and *w*_*OL*_=0.25 to the contributions corresponding to home, school, work, and “other locations”, correcting by reciprocity and determining a scaling factor in order that the NGM for each case leads to *R*_0_=1.5.

| POL-NL | POL-IT |
| --- | --- |
| $\begin{pmatrix} 2.95 & 0.72 & 1.81 & 0.56 \\ 0.72 & 2.86 & 1.69 & 0.48 \\ 1.81 & 1.69 & 2.39 & 0.99 \\ 0.56 & 0.48 & 0.99 & 2.21 \end{pmatrix}$ | $\begin{pmatrix} 2.54 & 0.84 & 1.63 & 0.9 \\ 0.84 & 3.45 & 1.77 & 0.74 \\ 1.63 & 1.77 & 2.22 & 1.17 \\ 0.90 & 0.74 & 1.17 & 1.43 \end{pmatrix}$ |
| POL-UK | SYN-ARG |
| $\begin{pmatrix} 3.26 & 1.49 & 1.87 & 0.73 \\ 1.49 & 3.60 & 1.82 & 0.80 \\ 1.87 & 1.82 & 2.03 & 1.08 \\ 0.73 & 0.80 & 1.08 & 1.69 \end{pmatrix}$ | $\begin{pmatrix} 2.61 & 0.98 & 1.53 & 0.93 \\ 0.98 & 3.48 & 1.73 & 0.57 \\ 1.53 & 1.73 & 2.50 & 0.86 \\ 0.93 & 0.57 & 0.86 & 1.14 \end{pmatrix}$ |

With the aim of exploring conditions under which our methodology would yield matrices with elevated contacts within the 15-30y age group, we examined scenarios characterized by a greater relative underreporting for this group. We considered scenario fU3, which is the same as fU2 but with inverted values for *f*_*U*1_ and *f*_*U*2_, and scenario fU4 where *f*_*U*1_ = *f*_*U*2_ = *f*_*U*3_/2. Table 7 displays the matrices obtained with our methodology for scenarios CM2-gf1-fU3 and CM2-gf1-fU4. We chose the CM2 structure since CM1 explicitly assumed in its construction that the 30-60y age group drove the contagion process. It is observed that, indeed, considering a higher relative underreporting for the 15-30y age group results in increased contacts for this group. Particularly, scenario CM2-gf1-fU4 produces matrices very similar to POL-IT and SYN-ARG. The overall agreement is remarkable, except for contacts involving the elderly group which was expected since the matrices in Table 6 do not account for the general reduction in contacts among older adults typical during the pandemic. While the matrices in Table 6 cannot be taken as reliable approximations to contact rates for our problem, considering all we have mentioned at the beginning of this section, it cannot be ruled out that some of the observed elements are part of a plausible scenario. On the other hand, the underreporting scenarios fU3 and fU4 (which we did not originally consider) are not implausible either. They could be explained by assuming, for example, that although the 15-30y age group probably exhibited more symptoms than children, there was less care and concern focused on them. We analyzed the control strategies from Section 3.2.2 for the new scenarios and found that for CM2-gf1-fU3, it is not possible to halt transmission by immunizing only group 3, whereas for CM2-gf1-fU4, it is possible but requires almost complete immunization of group 3 (95% for strategy (i) and 92% for (ii)). This is because a significant portion of transmission falls now within group 2. Therefore, the strategy of immunizing only group 3 would no longer be effective for these scenarios. Strategies (iii) and (iv), on the other hand, yield identical results to scenario CM2-gf1-fU1 presented in Table 5.

**Table 7:** Contact matrices *β*_*ij*_ (in units of *γ*) for epidemiological scenarios CM2-gf1-fU3 and CM2-gf1-fU4, where 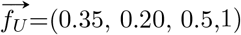 for fU3, and 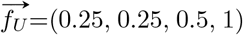 for fU4.

| CM2-gf1-fU3 | CM2-gf1-fU4 |
| --- | --- |
| $\begin{pmatrix} 1.85 & 0.92 & 0.92 & 0.20 \\ 0.92 & 4.24 & 1.64 & 0.40 \\ 0.92 & 1.64 & 2.32 & 0.80 \\ 0.20 & 0.40 & 0.80 & 0.00 \end{pmatrix}$ | $\begin{pmatrix} 2.72 & 1.36 & 1.36 & 0.21 \\ 1.36 & 3.72 & 1.57 & 0.42 \\ 1.36 & 1.57 & 2.57 & 0.85 \\ 0.21 & 0.42 & 0.85 & 0.00 \end{pmatrix}$ |

### 3.4 Discussion

Despite the uncertainties present in various aspects of the problem, the proposed methodology allowed us to obtain information about possible contact structures during the COVID-19 transmission process in the Greater Buenos Aires, and how these structures depend on the assumptions made to obtain them.

The assumptions underlying the contact structure CM1 appear to be reasonable, as for underreporting factors close to those given by fU1, plausible values of contact rates are obtained that are consistent with the formulated hypotheses. The CM2 structure also appears feasible, and when evaluating vaccination strategies, it is observed that in some aspects the results are similar to those obtained with the CM1 structure, while in other aspects, they are not. For example, the value of the fraction to be immunized, *φ*, which is necessary to stop transmission in different strategies, depends primarily on 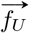 (observe the similarity of the values within each cell of Table 5). However, the differences between CM1 and CM2 have some notable consequences in the case of gf0 for control strategies where only the 30-60y group is immunized:

- In the cases of gf0-fU0 and gf0-fU1, it is not possible to stop transmission by immunizing only group 3 when assuming the CM1 contact structure, while it is possible with the CM2 structure.
- Conversely, in the case of gf0-fU2, the scenario with CM1 is more effective for that immunization strategy than CM2.

This can be understood based on the following considerations:

- For CM1, children only contact adults, so when they contribute little to transmission (cases gf0-fU0 and gf0-fU1), *β*_13_ is low. Since for this structure it was assumed that *β*_23_=*β*_13_, *β*_23_ is also low, which causes *β*_22_ to have a significant value as it has to account for incidence in 15-30y age group. As a consequence, immunization of group 3 has low impact on 15-30y incidence and is not able to stop transmission. This effect is not present in CM2 contact structure where *β*_23_ is a weighted average of the diagonal elements *β*_22_ and *β*_33_ and is not related to *β*_13_.
- An opposite effect is found when considering gf0-fU2 scenarios where the high under-reporting assumed for children leads to a significant contribution to transmission from them. This contribution is directly linked to group 3 in CM1 structure through a high *β*_13_ value, and so in this case, immunization of group 3 not only is enough to stop transmission, but is more effective than for CM2 structure, where there is a percentage of children that are not directly affected by immunization of group 3, since for CM2 *β*_11_*≠* 0.

These differences found between the CM1 and CM2 structures highlight the crucial role that social contact structures can play in some cases. In an emergency situation, such as the one experienced during the COVID-19 pandemic, the type of analysis conducted here could be used to detect and suggest specific aspects of the contact structure that are more important to investigate through field studies, such as conducting surveys.

Given the multiple uncertainties inherent in estimating contact matrices using survey data, as conducted in Section 3.3, the matrices obtained can only be considered as a possibility. However, the exercise proved useful because it prompted us to consider different scenarios for the originally conceived relative underreporting. Moreover, it highlights the importance of having a survey-based study of social contacts conducted under study-specific conditions to help narrow down the possible values of relative underreporting, which are always elusive to determine and, as we have seen, can have a decisive impact on the vaccination strategies to be implemented.

Based on the various explorations conducted, the CM2 structure emerges as an appealing candidate for the WAIFW matrix, as it exhibits great flexibility, capable of accommodating scenarios where transmission was focused on different age groups.

Even when compartmental age-structured models have been widely used during the COVID-19 pandemic[14, 37, 42], it is important to consider that when restriction measures are imposed, the implicit assumption of homogeneous mixing within each age group in Eq. (2) may not be valid. Particularly, under the conditions that existed in the Greater Buenos Aires during the pandemic, it is likely that a fraction of the population remained outside the contagion process. For example, those families who strictly adhered to safety regulations, with breadwinners performing virtual tasks. However, to the extent that the fraction of the population that participated in the contagion process had a similar age distribution to the total population, our results would not be affected.

## 4 Concluding remarks

In this work, we present a methodology that allows linking age-dependent transmission parameters with epidemiological data at the onset of the epidemic spread of an infectious disease. We applied it to the case of COVID-19 in Greater Buenos Aires, Argentina, in 2020, with the aim of obtaining information about the contact rates between different age groups based on knowledge of the fraction of reported incidence by age. Since we have incomplete knowledge of other quantities such as susceptibilities, infectivities, and relative underreporting factors by age group, we formulated hypotheses to estimate them, leading us to define various epidemiological scenarios. Despite the uncertainties present in the problem, our study provides information that could be useful in the event of another airborne emerging disease, where mobility restrictions similar to those in place during the first wave of COVID-19 in Greater Buenos Aires (GBA) were in effect.

For the age partition considered, we found that for the relative underreporting factors that seemed reasonable to us a priori (those discussed in Section 3.1.3: fU0, fU1, fU2), the 30-60y age group was the most active one. If a sterilizing vaccine had been available at that time, immunizing less than half of the 30-60y age group would have been sufficient to halt the epidemic in most of the analyzed scenarios. However, if a higher underreporting were to be observed in the 15-30y age group (as would be inferred by taking the contact matrices estimated in Section 3.3 as valid), then this group would bear a significant burden, and immunizing only the 30-60y age group would no longer be a convenient strategy.

On the other hand, we found that the data impose certain constraints on the hypotheses that are not always compatible. For example, it is not compatible with the epidemiological data to assume that children are less susceptible and infectious than adults and that they also have a much higher level of underreporting than adults. Both hypotheses together lead to an excessive level of activity among children, which is not consistent with the restrictions imposed on them (absence of classes and formal sports activities).

This type of study can provide an insight into where to focus complementary epidemiological studies at the beginning of an epidemic when there is a need to gather information quickly, and there is no time for extensive studies. For example, it can help focus serological studies on estimating the relative underreporting of specific age groups or direct the design of social contact studies to gather information on aspects suspected of playing a more relevant role.

## Data Availability

Data is available at: https://datos.gob.ar/dataset/salud-covid-19-casos-registrados-republica-argentina/archivo/salud fd657d02-a33a-498b-a91b-2ef1a68b8d16

## Acknowledgements

We acknowledge financial support from the Consejo Nacional de Investigaciones Científicas y Técnicas (CONICET) through PIP 0292 (2017-2019) and PIP 0266 (2022-2024). G.F. is member of the Scientific Career of Consejo Nacional de Investigaciones Científicas y Tecnológicas-CONICET (Argentina). S.S. acknowledges a fellowship from CONICET.

## Footnotes

1 Matrix *M* ^*′*^, defined below in eq.(11), is a nonnegative matrix, so it has a nonnegative eigenvalue 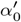 such that the modulus of all the other eigenvalues, 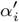, do not exceed 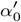, and to 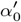 corresponds an eigenvector with nonnegative components (this is proved in ref. [25]). As 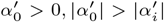 implies 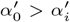. On the other hand, we have assumed that *α*_*D*_ is a dominant eigenvalue of *M* : *α*_*D*_ > *α*_*i*_, *∀i*. Since eigenvalues of *M* and *M* ^*′*^ are related by 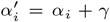 (and eigenvectors are the same), it follows that: 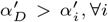, so 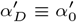, that is, it is the eigenvalue whose eigenvector, 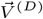, has all nonnegative components.

2 It is worth noting that when the value of the constant *c* obtained in each case is used to calculate the corresponding basic reproduction number, *R*_0_, for the unrestricted situation (i.e., *w*_*H*_ = *w*_*S*_ = *w*_*W*_ = *w*_*OL*_ = 1), values between 2.9 and 3.7 are obtained. These values represent plausible estimates for *R*_0_ during the COVID-19 epidemic in the absence of restrictions.

